# Quantitative EEG-Based Deep Learning for Neonatal Seizure Detection using Conv-LSTM

**DOI:** 10.1101/2025.11.04.25339496

**Authors:** Sonali Santhosh, Danilo Bernardo

## Abstract

Neonatal seizures cause significant morbidity and mortality, both acutely and in the long term, contributing to adverse neurodevelopmental outcomes. Traditional EEG seizure detection by human experts is constrained by limited efficiency, scalability, and objectivity, which can lead to delay diagnosis and hinder optimal outcomes. Deep learning methods have shown promise neonatal seizure detection, however, more recent DL architectures have not been robustly evaluated Here we evaluated a novel DL architecture Conv-LSTM in neonatal seizure detection. This study involved a publicly available dataset with 79 term neonates, in which 20 infants were randomly selected. The 20 EDF files were processed through bad signal removal, band-pass filtering, and fixed-length epoch segmentation. Feature extraction was then performed using Principal Component Analysis on both entropy-based and wavelet decomposition features, followed by standardization and temporal duplication. The model was able to universally qualify the neonates and achieve an overall accuracy of 86%. Out of all 20 patients, the model could accurately predict 90% of the non-seizure activity and 76% of the seizure activity. Thus, this study determines the potential of Conv-Bi-LSTMs in accurately detecting neonatal seizures based on EEG data, showing promise for reducing the mortality rates and negative long-term impacts on susceptible neonates.

## Introduction

Every year, up to 5.5 out of 1000 infants are affected by neonatal seizures; this number increases significantly among those born prematurely, leaving lifelong consequences, especially in infants who had a delay in diagnosis or treatment of seizures (Kraweic & Muzio, 2023). In an infant’s critical first moments—the first twenty-eight days of their life, known as the neonatal period—their brain is a whirlwind of activity, making it difficult to distinguish abnormal patterns like seizures, which can strike without warning and leave devastating effects. Seizures are defined as an abnormal discharge of neurons in the brain (Stafstrom & Carmont, 2015). For neonates specifically, seizures often occur due to hypoxic-ischemic encephalopathy (HIE), a birth complication that impedes or causes a lack of ample blood flow and or oxygen delivery to the brain (Allen & Brandon, 2011). Alternatively, they may also be caused by infection, hemorrhage, stroke, or additional etiologies (Kraweic & Muzio, 2023). Accurate diagnosis allows for these vulnerable patients to recover efficiently, however, the absence of timely detection of seizures leads to several extended detrimental effects. A study published by the American Academy of Pediatrics found that in a subset of 89 infants with neonatal seizures, over 35% experienced deleterious long-term impacts, such as death or severe cognitive disabilities caused by permanent brain damage (Stieren et al., 2024).

Electroencephalogram (EEG) data is commonly used by neurologists to diagnose seizure activity in the general population, including neonates. By placing a differing number of electrodes on the patient’s scalp, medical professionals are able to non-invasively measure the electrical activity in clusters of neurons throughout the brain (Light et al., 2010). EEG is proven to be the most effective modality to diagnose seizures, specifically continuous EEG (cEEG) recordings, which are essentially uninterrupted EEG recordings at any time from a few hours to a few days straight. This prolonged EEG data allows for the most precise diagnosis of patients who are at high risk for seizures or epileptic syndromes. Glass et al. conducted a study involving 426 neonates with suspected seizures, monitoring them with cEEG for a minimum of 24 hours. Their findings revealed that over 59% of the infants experienced more than seven seizures, emphasizing the high seizure burden in this population and the critical role of cEEG in accurate seizure diagnosis and management (Glass et al., 2017). If facilities are unable to support the high demand of cEEG technologies, they may turn to amplitude-integrated EEG (aEEG), which involves a simpler methodology and a reduced number of channels (1-2), streamlining the overall diagnosis process. However, it is strongly recommended to use cEEG if available, as aEEG is both less specific and less sensitive to diagnosing seizures than the former (Kim et al., 2022).

The challenge, however, arises from the lack of clear clinical manifestations of seizures in neonates compared to those frequently seen in older age groups. Since most observable symptoms in neonates are subtle and nonspecific, diagnosing neonatal seizures becomes especially challenging. Additionally, infants may exhibit atypical seizure behaviors, namely apnea or abnormal movements of the eyes, tongue, or mouth, leading to the mistaken assumption that the infant is clinically stable (Neonatal Seizure Registry). Furthermore, prior studies have shown that the majority of neonatal seizures “are electrographic-only events without clinical symptoms,” reinstating the former issue (Kim et al., 2022). As a result, EEG recording becomes essential for accurate diagnosis. However, manual interpretation of neonatal EEG data does require significant expertise and time. As noted by Tang & Zhao, the immature brain’s physiology differs highly from older age brackets, making seizure detection and classification less straightforward (Tang & Zhao, 2024). This requires the most highly qualified, trained pediatric neurologists to be present to accurately diagnose seizures in infants, and many healthcare facilities may lack access to such specialized expertise, particularly in resource-limited settings (Rennie et al., 2019). Regardless, the overall process of neonatal seizure detection tends to be complex and resource-intensive, making it not only time-consuming but expensive as well.

ML offers healthcare providers to significantly enhance accuracy in their diagnosis, personalize individual treatment plans, and forecast specific patient outcomes (Sim et al., 2021). In Neonatology specifically, ML has been implemented in a multitude of ways, such as being able to monitor vital signs, detect abnormalities in electrocardiogram (ECG) and EEG data, and aid personalized infant care. Additionally, predictive analytics have allowed for efficiency in anticipating complications such as respiratory disease, jaundice, infections, and seizures (Chioma et al., 2023). Currently, ML serves a supplementary role in healthcare due to the large limitations still present in fully incorporating AI in clinical practices. For instance, a lack of large datasets that can be used to train these models on specific conditions. However, as AI technology advances, its influence on direct patient care and potential to improve diagnostics will be significant (Chioma et al., 2023).

Building on the challenges outlined in neonatal seizure diagnosis, the former limitations have driven the adoption of more sophisticated techniques—automated models. Many traditional Machine Learning models, such as Support Vector Machines (SVMs), Random Forest (RF), and conventional Neural Networks, have been studied with fairly high accuracy. Among neural networks, Convolutional Neural Networks (CNNs) are widely recognized for extracting spatial patterns, such as those within EEG signals, Deep Neural Networks (DNNs) leverage multiple hidden layers for immense feature learning, and Recurrent Neural Networks (RNNs) are typically preferred for sequential data. For instance, Debelo et al. utilized a CNN to classify neonatal seizures, achieving 88.6% accuracy in multiclassification (Debelo et al., 2023). Zeedan et al. proposed a Long Short-Term Memory (LSTM) network, a specific type of RNN, and achieved 87.7% accuracy in neonatal seizure detection. In a separate study, Alharthi et al. achieved approximately 95% accuracy using a Conv-Bi-LSTM to detect general epilepsy from EEG signals, underscoring the hybrid model’s promise. Despite its compelling performance, the Conv-Bi-LSTM has not yet been applied specifically to neonatal seizure detection.

However, there is a limited exploration of hybrid models on neonatal seizures, especially a Convolutional Bidirectional Long Short-Term Memory (Conv-Bi-LSTM) model, which effectively analyzes both spatial and temporal data from EEG signals. Conv-Bi-LSTMs have been proven to be particularly valuable for general seizure detection due to their enhanced feature extraction capabilities, but have yet to be applied specifically to neonatal EEG data. Given that neonatal seizures are associated with a mortality rate of approximately 10%, with 50% of the survivors suffering from significant disabilities, advancing seizure diagnostic methods is crucial (Rennie et al., 2019). Thus, this study aims to transform neonatal care, reducing mortality and morbidity rates worldwide, by addressing the following question: How effectively can a Conv-Bi-LSTM model detect seizure events in neonates by learning both spatial and temporal patterns from multi-scale EEG features in clinician-annotated data, and how successful is its performance across varying patient conditions?

### Literature Review

#### Methodology

This study utilized secondary collected EEG data from the Neonatal Intensive Care Unit (NICU) at Helsinki University Hospital in Finland, which is publicly available on Zenodo, a site designated to share open-source data (Stevenson et al., 2018). The multi-channel EEG data was collected from 79 neonates prone to seizures with various clinical backgrounds. From the seventy-nine infants, this study randomly selected twenty patients for analysis, using a random numerical generator to prevent any training biases. The EEG files were recorded through the NicOne EEG amplifier at a sampling frequency of 256 Hz, using the standard 10-20 EEG montage depicted below in Figure 1. An average recording duration of 74 minutes with a vast interquartile range of 64 to 96 minutes was present in the data. Each infant’s file was annotated for seizures by 3 experts with over ten years of experience in neonatal EEG using the Nicolet Reader software. The data was marked as seizure activity if, and only if, the distinguishable electrographic event was atypical with continuous “repetitive evolving spike/sharp waves or rhythmic waveforms” and consisted of a distinct beginning and end. Each specialist recorded an average of 460 seizures across the database, alongside differential seizure activity, with 39 neonates having seizures and 22 neonates classified as seizure-free unanimously. The database consists of 79 EDF files, the most commonly used file format for EEG, a CSV file containing each patient’s clinical information and background, and 3 CSV files labeled ‘A’, ‘B’, and ‘C’ with each expert’s individualized annotations.

**Figure 1.**
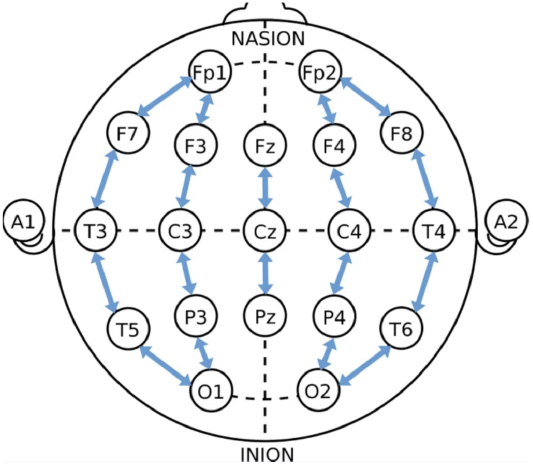
EEG Montage used in collecting data from all 79 neonates across 19 EEG electrodes, 1 ECG channel, and 1 Respiratory Effort channel. (Visual was created by Stevenson et. al, 2018). https://doi.org/10.1038/sdata.2019.39

Various preprocessing measures were executed to ensure high-quality EEG signals within the data, including noise removal, band-pass filtration, and data segmentation. Within the twenty-one channels available per patient in the Helsinki dataset, two of those channels are Electrocardiogram (ECG) and Respiratory Effort Channels. Although multimodal data fusion may be useful in some cases, this study chose not to incorporate the corresponding two channels to focus on EEG analysis solely and prevent any cross-channel contamination. Without the removal of the two channels, the additional data may disrupt the model from learning the patterns within the EEG, specifically. In order for the model to focus on key signals within the neonatal EEG, the raw data were band-passed filtered between 0.5 to 30 Hz, the ideal range to capture infantile frequencies (Lundy et al., 2023). Independent Component Analysis (ICA) was later directly applied to the filtered EEG data of a single patient to explore its potential for artifact removal and data cleaning. However, the process was found to be time-consuming, and automating ICA for a broader patient scale would have required significant manual intervention, making it impractical within the constraints of the study.

Subsequently, the data is segmented into non-overlapping fixed-length windows of one second each to correspond with the annotations. Initially, the model was evaluated based on the annotation approach of majority voting (≥2 expert agreements). However, the model yielded more accurate results with a more inclusive annotation approach, with seizure events being classified if at least one expert identified them (Figure 2). Consequently, instead of requiring the consensus of two or more experts in labeling an individual’s second of data as ‘1’ or seizure activity, it only requires one expert to have identified the data as seizure activity to label it as such. Otherwise, the data is simply labeled ‘0’ as normal activity.

**Figure 2.**
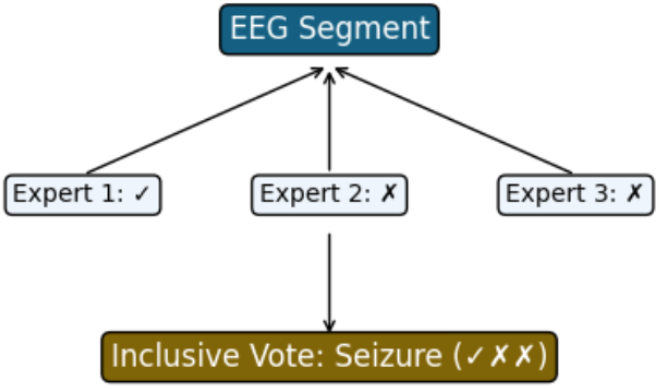
The annotation selection process used in the study. (Visual generated by student researcher through Canva, 2025.)

Several feature extraction methods were investigated in this study. The features that yielded the most optimal performance were wavelet transformation and entropy-based features. Unlike the Fourier transform, wavelet transformation is able to recognize time-varying frequencies and decompose and capture the signal into different components. Specifically, Discrete Wavelet Transformation (DWT) was used in this study to conduct scaling and wavelet functions to offset the initial scaling (Gosala et al., 2023). Regarding feature extraction, the Daubechies-4 (db4) wavelet was selected for its improved performance on biomedical signals such as EEG. A breakdown of the DWT decomposition formula is displayed below in Figure 3. From each decomposition level, mean, measuring central tendency; standard deviation, collecting the signal’s variability; and energy, capturing the signal’s power, were extracted.

**Figure 3.**
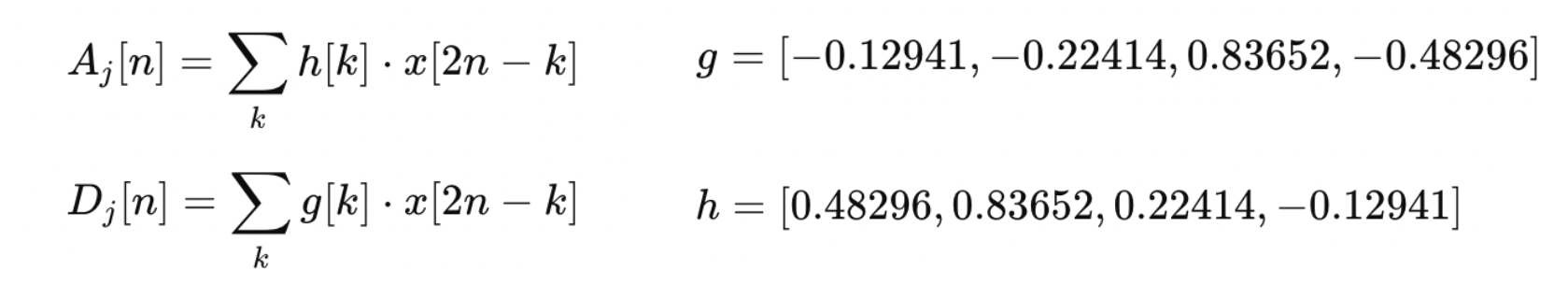
Formula breakdown for Daubechies-4 wavelet. (Figure for formulas created by student researcher through Canva, 2025).

Additionally, entropy-based features were also extracted. Entropy measures the randomness or instability of the EEG signals, which is useful for quantifying the amount of information present in the signal. Both Sample Entropy (SampEn) and Permutational Entropy (PermEn) were extracted and concatenated into a single matrix. SampEn, calculated below in Figure 4, is used to estimate the complexity and predictability of an EEG signal by measuring the likelihood that a sequence of signals remains similar when extending or de-extending the length (Dastgoshadeh & Rabiei, 2023). PermEn, also calculated below in Figure 4, extracts the order patterns of a signal to calculate complexity instead of simply magnitude, essentially evaluating the randomness of local sequences within the signal. All five features are extracted from each one-second epoch for each patient and then embedded into a complete feature matrix to undergo further data augmentation before model training.

**Figure 4.**
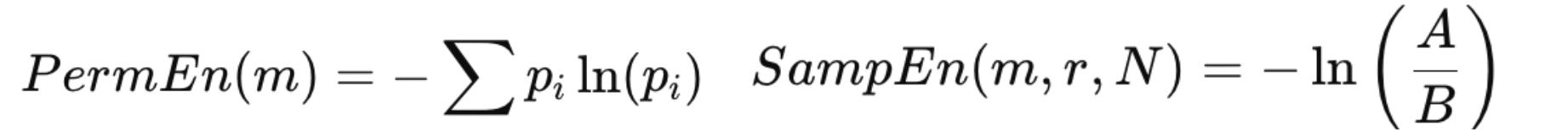
Formulas for entropy-based features: Permutational and Sample Entropy. (Figure for formulas created by student researcher through Canva, 2025).

Computational constraints are a common limitation in machine learning, and at this stage, the dimensionality of the combined feature set has become increasingly large across the twenty patients. To address this, Principal Component Analysis (PCA) was implemented to reduce redundancy and increase efficiency. PCA limits the data size to a certain number of feature components that capture the specified percentage of the data variance. Initially, PCA was configured to explain 95% of the variance. However, when training across multiple patients, where each infant’s data size varies, it became necessary to set PCA to a fixed number of components (N=160) to maintain consistency when concatenating data.

This study proposed a two-part Hybrid model, Conv-Bi-LSTM, for its distinct advantage of analyzing spatial and temporal time series data. The model integrates a Convolutional Neural Network (CNN) for feature extraction due to its prowess in visual image analysis, proving it useful for EEG signal selection (Rakhmatulin et al., 2024). A CNN utilizes multiple convolutional layers to gather data on grid-like features, identifying patterns and learning directly from the data. However, a CNN is unable to account for time series data and cannot gauge sequential information from EEG signals that is useful for accurate detection. The second component of the Hybrid model, a Recurrent Neural Network (RNN), comes into play here. An RNN is, once again, a superset of Neural Networks, with a specialty in introducing a dimensionality of time to data processing. The RNN uses feedback loops to “feed back” every output acquired as input alongside the additional input from every time step. This process allows the model to retain information from previous time steps and update its memory accordingly. This aspect makes the RNN model well-suited for tasks that require models to understand patterns across a long period of time, such as the EEG task in this study (Das et al., 2023). That being said, an RNN is known to undergo a particular issue. During backpropagation, an algorithm used to train neural networks, the RNN is affected by the vanishing gradient problem, in which the gradients used to adjust the model ultimately vanish during large-value data training (Tam et al., 2023). A Long Short-Term Memory (LSTM) Model, a subcategory of RNNs, addresses this limitation by having “nonlinear, data-dependent controls” within its cells that prevent the size of the gradients from getting increasingly smaller (Sherstinsky, 2020). A Bidirectional LSTM (Bi-LSTM), subsequently, contains both forward and backward-facing hidden layers, allowing the model to access greater data (Tam et al., 2023). Employing features of both a CNN and a Bi-LSTM ensures that the model is tailored to handle the unique challenges of analyzing both spatial and time-series data, making it a powerful solution for accurately interpreting EEG signals and identifying neonatal seizure activity.

The hybrid architecture used was altered several times during the course of this study. The following architecture, depicted in Figure 5, however, proved to be the most successful in performance. The model initialized through the Keras Sequential Model is renowned for its simplicity in stacking multiple layers (Chollet, 2015). For proper feature extraction, the model began with a convolutional layer with 64 units and a kernel size of three, with a Rectified Linear Unit (ReLu) activation function, frequently used with CNNs (Ide & Kurita, 2017), followed by another convolutional layer of 32 units, kernel size three, and ReLU activation. Subsequently, a Bi-LSTM layer with 128 units and an activation function of TanH was added to introduce non-linearity (Vijayaprabakaran & Sathiyamurthy, 2020). Finally, three dense layers of decreasing size (32, 16, and 1 unit (s)) and ReLU activation functions were appended for binary classification of the data. Aside from the former layers, Dropout, Batch Normalization, and L2 Regularization were used to prevent overfitting. Overfitting is when the model memorizes the training data and is unable to generalize to unseen data, such as the validation data, causing the model to underperform. Dropout is used to randomly “drop” a specific percentage (30% in this study) of nodes and connections after each step of the model. This allows the model to be readjusted on a new set of neurons to improve the model’s learning abilities. Batch Normalization normalizes the outputs of the first layer before inputting them into the next, creating faster convergence. L2 Regularization or Ridge Regularization enables a penalty on the loss function, which discourages large weight values, encouraging the model to become simpler and allowing for greater generalizability (Gygi et al., 2023).

**Figure 5.**
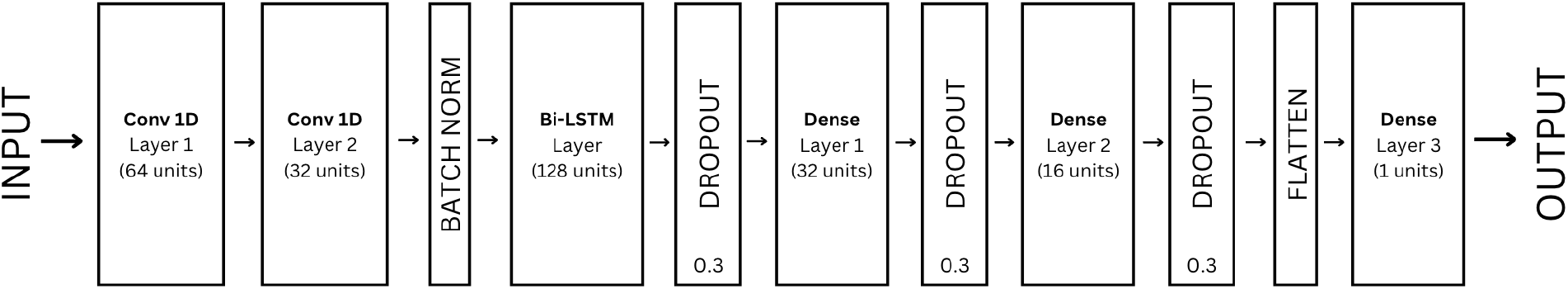
The following visual represents the architecture that the model utilized. (Visual generated by student researcher through Canva, 2025.)

The training data with a size of (195802, 3, 160), referring to 195,802 EEG samples across the twenty patients, 3 total timesteps, and 160 features per timestep, was then split into a seven-to-three ratio for training and testing data, leading to a final training and testing data with sizes of (137061, 3, 160) and (58741, 3, 160), respectively. The model was trained on the Adaptive Moment Estimation (ADAM) optimizer, with a learning rate of 0.01. An optimizer functions as the power source behind the model, driving it to consistently improve its parameters during training, reducing the chance of any errors. ADAM is widely implemented in ML over other optimizers due to its efficiency and adaptability with large datasets (Wei, 2024). The data was then trained through the model using the Binary Cross-Entropy Loss Function for a total of ten epochs, representing ten passes of the Conv-Bi-LSTM model through the entire training dataset.

Multiple performance-based metrics must be recorded to best understand the efficiency of the model. A confusion matrix generates key results of the true positive (TP), true negative (TN), false positive (FP), and false negative (FN) rates of the model, indicating the sensitivity and specificity of this study. Sensitivity refers to the percentage of positive cases correctly identified by the model, or the TP score divided by the sum of the total number of TPs and FNs. Specificity, on the other hand, represents the percentage of negative cases correctly identified by the model, or the TN score divided by the sum of the total number of TNs and FPs. In this study, accuracy, precision, recall, and F1-score were also calculated through a traditional classification report. Precision explains a counterpoint to the previously mentioned metric of Sensitivity, also known as recall, as it divides the TP score by the sum of both the TP and FP scores instead. Precision, thus, depicts the percentage of positive predictions accurately made by the model. Finally, the F1-score is the harmonic mean of both the Precision and Recall/Sensitivity scores (Rainio et al., 2024). All experimental procedures, including coding, were done in the Google Colab Jupyter Notebook software for easy editing.

## Results

The Hybrid Conv-Bi-LSTM model achieved a global accuracy of 86% across the entirety of the combined testing dataset containing 20 neonates. This represents the success of the model in distinguishing seizure and normal brain activity within infants suspected of seizures by analyzing their EEG data itself. On average, the model had relatively similar Precision and Recall rates for both seizure and normal states, as described in Figure 6. The Receiver Operating Characteristic (ROC) Curve in Figure 7 demonstrates that although the model performed more effectively than random chance (0.5 Area Under the Curve (AUC)), with an area of 0.907, there is still room for improvement regarding the model’s tendency to misclassify seizures with less distinct patterns.

**Figure 6.**
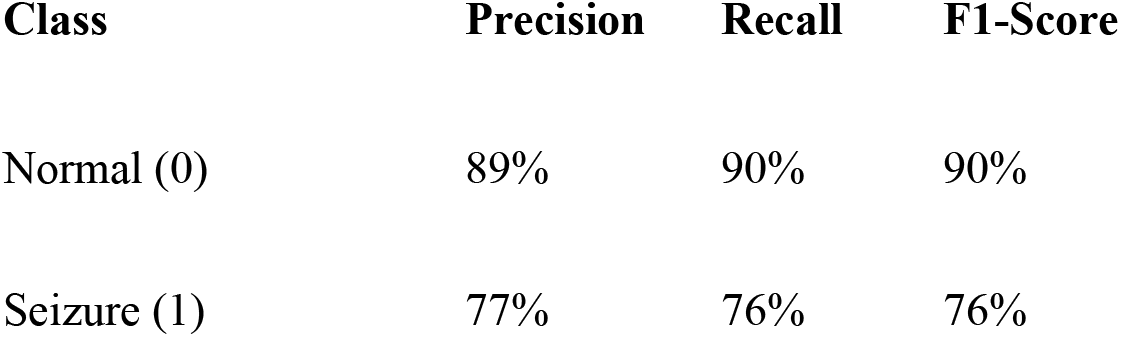
Classification report for the Conv-Bi-LSTM model depicting Precision, Recall, and F1-Score for both normal and seizure classes. (Table created by student researcher through Google Docs, 2025.).

**Figure 7.**
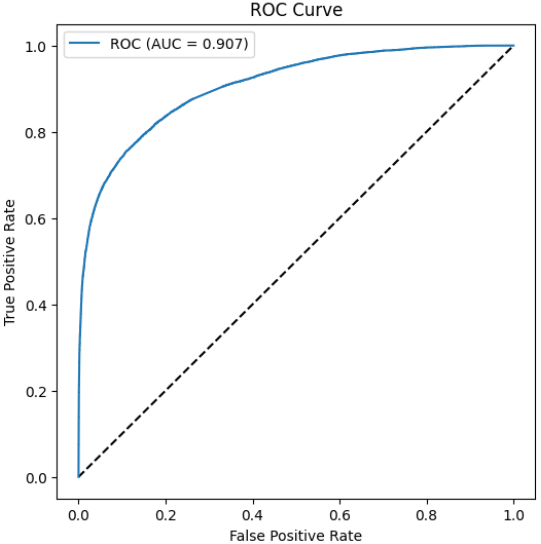
Receiver Operator Characteristic (ROC) Curve illustrating tradeoffs between True Positives and False Positives produced by the model. (Graph generated by student researcher through Google Colab, 2025.)

A more detailed breakdown of the results is displayed in the Confusion Matrix below in Figure 8. The matrix reinforces the model’s high TN rate while simultaneously minimizing both FNs and FPs. Additionally, the clear class imbalance between normal and seizure states is visible within the figure, helping justify why the model was prone to favor negative cases. This imbalance likely contributed to the model’s difficulty in identifying seizure epochs, as the model was unable to train over equal amounts of data from both classes, being significantly more exposed to normal data. Unfortunately, in any EEG dataset collected from a real clinical setting such as the Helsinki NICU, there will be a distinct class imbalance, as even seizure-prone infants will only be seizing for a fraction of the time they are being monitored. Despite this, the Conv-Bi-LSTM was able to achieve a reasonable trade-off between sensitivity and specificity, visible within the evaluation metrics presented in Figure 6.

**Figure 8.**
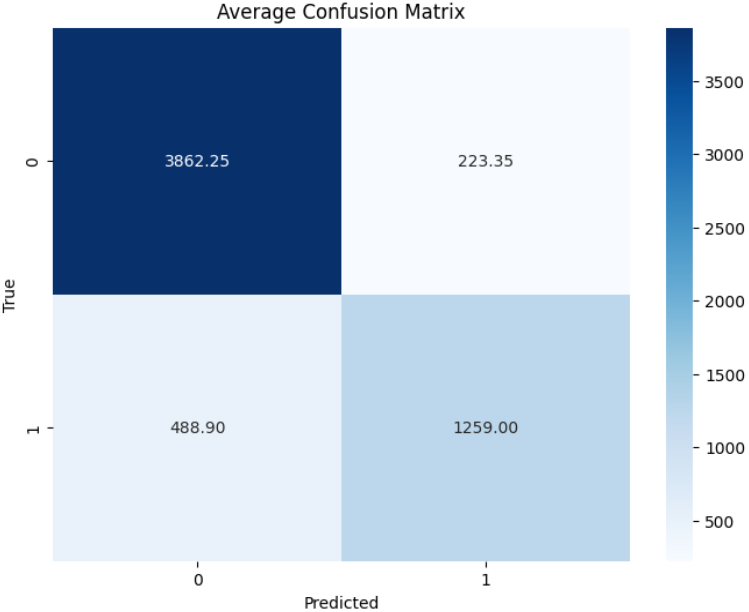
The confusion matrix shows four outcomes: false negatives (FN, missed seizures), true positives (TP, correct seizure detection), true negatives (TN, correct normal detection), and false positives (FP, normal data misclassified.) (Visual generated by student researcher through Google Colab, 2025.)

Additionally, the loss and accuracy graphs shown below (Figures 9 and 10), relay key indicators of a well-performing model. In Figure 9, both the training and validation loss is shown to be consistently decreasing after running through the ten epochs, indicating that the model was able to actively understand the underlying patterns present within the data. Figure 10 supports this by showing a similar minimal gap between training and validation accuracy as they both increase steadily through the ten epochs. Instead of overfitting to random noise or other irrelevant details, the Conv-Bi-LSTM generalized to unseen data, accurately detecting seizures in neonates.

**Figure 9.**
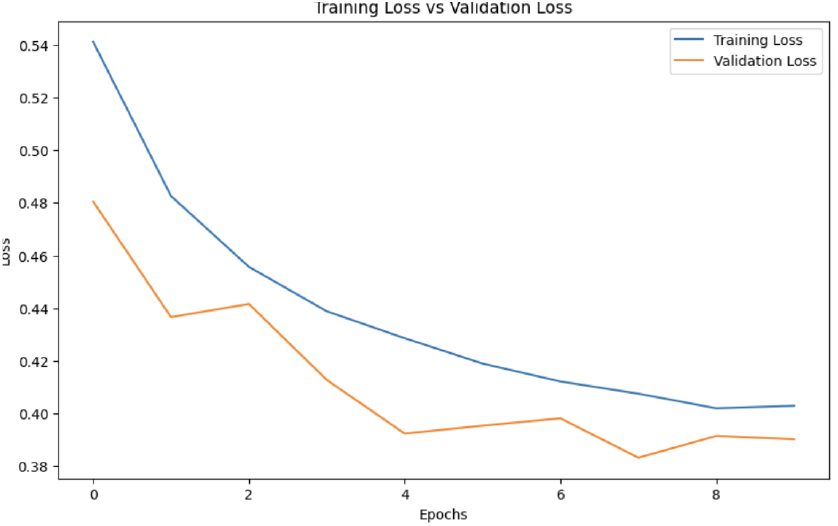
Displays the training vs. validation loss across multiple epochs. (Graph generated by student researcher through Google Colab, 2025.)

**Figure 10.**
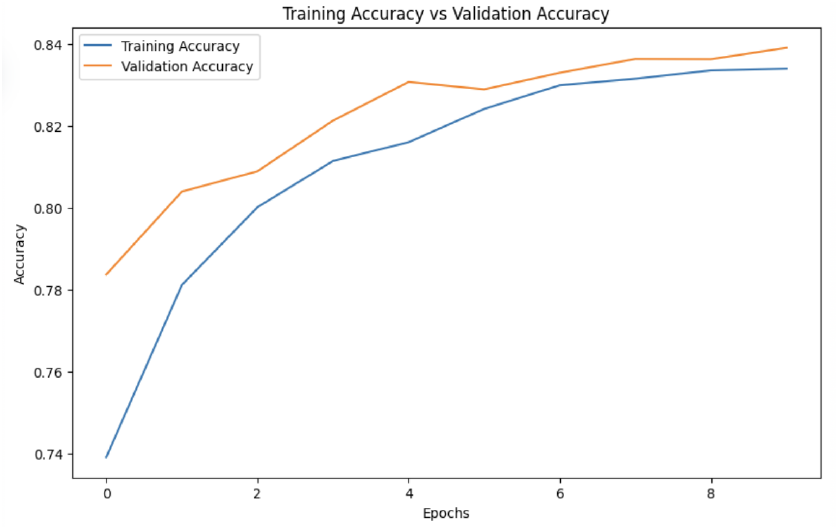
Displays the training vs. validation accuracy across multiple epochs. (Graph generated by student through Colab, 2025.)

Although viewing results of the combined dataset is essential, it is equally important to analyze patient-by-patient performance. Combined metrics often conceal important variations between cases, hiding deeper insights into the model’s achievement. Hence, in this study, after training the Conv-Bi-LSTM model across 70% of the combined dataset, the model was tested on the untrained 30% and individually evaluated across the 20 neonates as well, highlighting strengths, inconsistencies, and unique patterns in the EEG data for each patient. Across all 20 patients separately, the model achieved a mean per-patient accuracy of 87.79% with a standard deviation of 7.73%. By isolating each case specifically, it became easier to identify the model’s strengths and limitations. Infants with clear seizure patterns and distinct features were detected consistently, while more subtle or overlapping EEG patterns faced greater challenges, going undiscovered or being falsely identified as seizures.

A Pearson Correlation Coefficient (PCC) analysis was then performed to identify which extracted EEG features were most significant in improving the model’s performance. A PCC test statistically evaluates both the strength and direction of a linearly correlated relationship between two distinct variables, in this case, EEG features (X) and seizure detection accuracy (Y). This study relies on a critical assumption, that such a linear relationship exists between the variables that are being analyzed. While alternative statistical tests do assess non-linear relationships, for example, Spearman’s correlation test, the PCC test was selected instead for its simplicity and interpretability. A mathematical breakdown of this method is shown in Figure 11 below.

**Figure 11.**
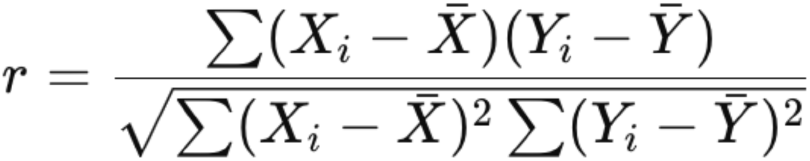
The formula for the Pearson Correlation Coefficient between EEG features (X) and seizure detection accuracy (Y) determines the strength and direction of the correlational relationship. (Formula was created by Dhanakotti, 2024.) https://dkaarthick.medium.com/correlation-pearson-vs-spearman-rank-correlation-dc3e5420339b

A PCC test value may range from -1 to 1. A positive value indicates a direct relationship between the two variables, such that as the feature’s value increases, so does the model’s predictive accuracy. A negative value implies the opposite, as a feature’s value decreases, the model’s predictive accuracy increases instead. Figure 12 reveals the overall distribution of correlations across all the extracted features, from both wavelet decomposition and entropy-based features. A majority of features had a positive correlation of 62.3% for seizure detection accuracy, although there was still a significant portion of features that had a negative correlation of 37.7%, reflecting a diverse set of features.

**Figure 12.**
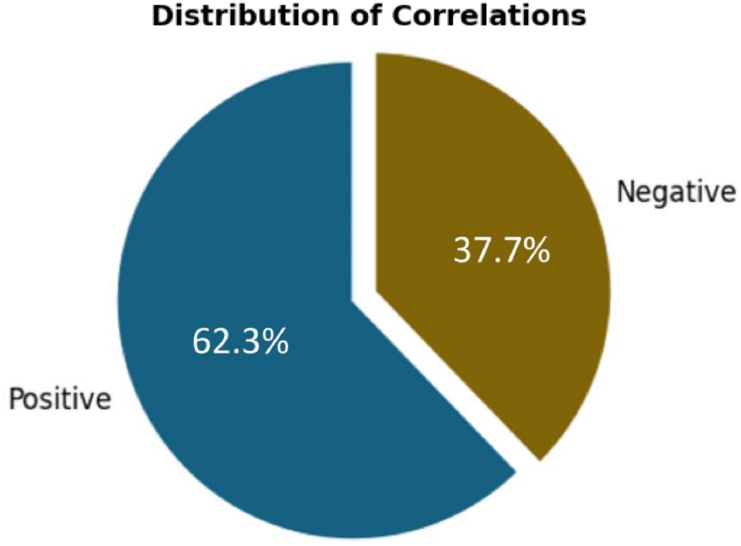
The distribution of correlations represents the amount of positively correlating features and negatively correlating features across the extracted selection. (Graph generated by student researcher through Google Colab, 2025.)

Figure 13, however, expresses the correlation strength of the same features across all 20 patients. An absolute PCC value closer to one signifies a stronger correlation between the variables, while a value closer to zero suggests weak or no linear correlation. This analysis not only uncovered which extracted features most strongly correlated with successful seizure classification but also highlighted a critical insight: no single feature reliably detected seizures to the same correlational strength in every patient. Each infant had unique seizure biomarkers explaining why some individuals posed greater classification challenges for the model than others, supported by the wide model performance variability within the accuracy range of 72% to 99% across all twenty neonates. These findings emphasize the need for personalized patient analysis for improving seizure detection outcomes.

**Figure 13.**
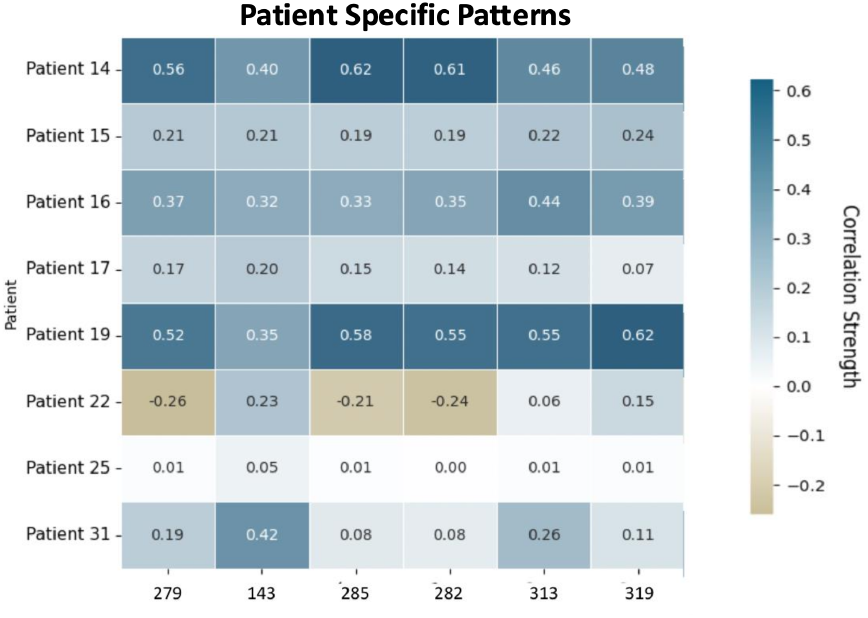
Displays different individual correlations for the same features across each patient, highlighting the variability in detection ability. (Graph generated by student researcher through Google Colab, 2025.)

## Discussion

The most notable outcome of this analysis was the recognition of multi-feature training as essential in neonatal seizure detection. Since this study used multiple extracted features within entropy-based and wavelet decomposition domains, the Conv-Bi-LSTM model maintained a global accuracy of 86%. However, variability was present on a patient-by-patient basis, with a range in performance from 72% to 99% in all twenty neonates. Despite this, Patient 6 (labelled in the Helsinki dataset as eeg22.edf), who achieved the 72% accuracy, still retained a reasonable validity, indicating a baseline consistency present within the model. This result confirms the model’s ability to differentiate between seizure and normal EEG events effectively. When analyzing both Patient 6 and Patient 19’s (labelled in the Helsinki dataset as eeg62.edf and the top-most performing patient at 99% accuracy) EEG data, key insights into the model’s pattern prediction ability were discovered. Sharp transient patterns with rhythmic discharges and frequency shifts within Patient 19 and other similarly-performing patients helped the model achieve the strong results. The model appeared to identify significant EEG features while struggling with more intermediary patterns within inconclusive or difficult-to-interpret data.

Compared to the prior models discussed, the hybrid Conv-Bi-LSTM performed quite similarly. However, it is essential to address that having equivalent access to resources and collaborating with larger research teams, comparable to other studies, could significantly optimize the model’s performance. Due to the independent nature of this study and restricted access to advanced resources typical of large research institutions, this study ran into several limitations.

Primarily, this model was only trained and tested on twenty neonates, affecting its ability to generalize confidently to a larger population. Although the Helsinki dataset did contain a total of 79 infants suspected of seizures, as a consequence of the limited time and computational ability of the device, this study was conducted on a reasonable number of infants, which was determined to be 20. By testing this model and feature extraction method on a larger scale of data or an alternative dataset, it may improve the overall results and support clinicians’ confidence in the AI’s ability.

Second, a key limitation lies in the uncertainty of the annotations located in the CSV files of the dataset itself. Even expert neurologists and other professionals trained on reading seizures in EEGs for decades struggled to perfectly align with one another. Three experts were used in the Helsinki dataset to annotate individual seconds of each patient’s EEG, and correspondingly label the second as either normal brain activity or seizure activity. However, when reading each expert’s CSV file of annotations, clear distinctions between the labeling were visible, introducing variability to the model’s training process. By utilizing an inclusive training approach, efforts were made to better handle the present class imbalance. However, this method means that if only one expert labeled normal EEG activity as a seizure, the model was trained to treat it as a positive class ‘1.’ Consequently, if the model later predicted a similar case as a ‘1,’ it would appear as a TP but could be a FP, and the same issue applies in reverse. Unfortunately, it is unlikely that there would be any way to prove that annotations are completely valid, since neonatal seizure detection on EEG is a challenging task.

Finally, during the PCC analysis, each feature used in the correlational analysis was labeled with an attached number (Figure 13, e.g., 313, 194, etc.). Although the analysis explains the strength and direction of each feature, it does not inform the feature’s location or where exactly the feature was extracted from. For instance, Feature 143 may be extracted from the standard deviation of a specific decomposition level using the Db4 Wavelet analysis, and Feature 177 could be a result of Permutation Entropy from the entropy-based features extraction, but without explicit labeling, it is difficult to determine and precisely trace back each feature’s origin for deeper interpretation.

There are several life-changing implications associated with this study. Faster intervention and a severe reduction of long-term neurological damage are essential outcomes. Due to the universal adaptability of the model, it demonstrated the ability to offer personalized neonatal seizure detection. However, it is critical to recognize that this model and subsequent models are not intended to replace physicians in general, but offer as a complement to their diagnostic abilities. This model would especially be valuable in low-resource settings with higher infant mortality rates. In areas lacking 24/7 neurologist coverage, AI-based alternatives such as this model could serve as an essential back-up system, enhancing early intervention.

To strengthen future research, further data processing, feature optimization, data augmentation, along with model refinement are imperative. Although the current performance is successful, the model needs to retain its pattern detection ability on even the most demanding cases. Additionally, before proceeding toward additional areas for improvement, such as clinical testing, achieving at least 95% detection accuracy is vital to ensure efficiency. Thereafter, future directions should be focused on attaining real-world validation through clinical integration to reinforce and examine the model’s reliability. Ultimately, to improve this technology’s accessibility, Edge AI deployment should be investigated to better aid low-income settings by offering the AI model on the “edge” of the network, providing real-time decision making (Meuser et al., 2024). This may permit the model to function on portable devices and proceed without consistent internet access or expensive infrastructure, thereby expanding its reach toward under-resourced hospitals and medical facilities in need of improved neonatal care.

## Conclusion

This study explored the effectiveness of a Convolutional Bidirectional Long Short-Term Memory Network (Conv-Bi-LSTM) in detecting seizures through neonatal EEG data. The results of this research aimed to ease pediatric neurologists’ ability to detect neonatal seizures efficiently and improve patient outcomes by reducing infant mortality and the long-term negative effects of undetected seizures within neonates.

To enhance infant seizure detection, the model was trained on entropy-based and wavelet decomposition features, resulting in a strong overall performance. Working as a universal classifier, the model obtained a global accuracy of 86%, with 90% specificity and 76% sensitivity across twenty randomly sampled neonates. These results demonstrate the strong capability and potential for hybrid architectures such as the Conv-Bi-LSTM in detecting neonatal seizures consistently across a diverse patient range. Unfortunately, despite the promising outcomes, due to the absence of large-scale team resources, the model was developed under limited data, affecting its reliability in generalization across the neonatal population. However, through this research, the unique nature of feature biomarkers was discovered, emphasizing the need for detailed patient analysis, as specific features resulted in different correlations towards individual patients.

Therefore, this study not only offers a feasible, interpretable model for early seizure detection but also advances the continuously growing field of AI in neonatology and patient care. With future improvements in the model’s sensitivity and clinical integration, the Conv-Bi-LSTM model holds promise to function as a crucial support system to increase accessibility in low-income hospitals, improving health outcomes for vulnerable infants worldwide.

## Data Availability

All data are available online at https://zenodo.org/records/1280684

